# Correlations between individual factors, meteorological factors, and hemorrhagic fever with renal syndrome in the Tai’an area of China, 2005–2019

**DOI:** 10.1101/2020.07.11.20151373

**Authors:** XiuJuan Bi, Shuying Yi, Aihua Zhang, Zhenghua Zhao, Yunqiang Liu, Tao Wang, Chao Zhang, Zhen Ye

## Abstract

Hemorrhagic fever with renal syndrome (HFRS), is a serious threat to human health. The relative risks factors for different occupations, ages, and sexes are unknown.

The results showed that compared with the whole population, the risk ratio was 5.05 (p <0.05) among the rural medical staff. GAM showed that air temperature was positively correlated with disease risk from January to June and that relative humidity was negatively correlated with risk from July to December. From January to June, the cumulative risk of disease increased at low temperatures.

Rural medical staff showed a high risk of developing the disease. The possibility of human-to-human transmission of HFRS among rural medical staff is worthy of interest and deserves to be explored by further studies. Moreover, air temperature and relative humidity are important factors that affect the occurrence of the disease. These associations show lagged effects and differing effects according to the season.

## Introduction

HFRS is caused by various types of hantavirus and is a natural epidemic disease with rodents as the main source of infection(1). Hantavirus is an important zoonotic pathogen found on all continents except for Antarctica and is associated with HFRS and hantavirus pulmonary syndrome(2, 3).

The main host animals are rodents, followed by cats, pigs, dogs, etc. *Apodemus agrarius* and *Mus norvegicus* are the main hosts and sources of infection in China. Raising pet rats or keeping rats can also cause Seoul virus infection because of close contact, further leading to the occurrence of HFRS(4-7). The main routes of transmission include contact with rodent feces as well as respiratory tract, digestive tract, and contact transmission.

The Seoul virus has spread globally and has been found in mice in the UK, France, Sweden, and Belgium(8). The disease is prevalent worldwide, with Eurasia being the main area of disease(9) and China being a more serious area(10). In 2000–2017, Russia reported 131,590 cases of viral HFRS caused by six different hantaviruses(11). The average annual incidence rates were 6 cases per 100,000 people in western Russia and 0.4 cases per 100,000 people in eastern Russia. From January 2014 to June 2019, nearly 60,000 cases of HFRS occurred in mainland China, with 360 deaths and a case fatality rate of 0.6% (data source, Chinese CDC http://www.chinacdc.cn/). Among regions in China with HFRS, the areas with the highest prevalence include Northeast China, Shanxi, and Shandong Provinces(12). Tai’an is a prefecture-level city in Shandong Province, with incidence differing from that in Shandong as a whole. Many studies have reported that the main group of patients is farmers(13, 14). It is not clear whether there are groups with higher risks of disease than farmers. Moreover, whether there is human-to-human transmission of HFRS as an infectious disease has not been reported. In addition, the differences in risks according to age and sex are not clear. In addition to being related to hantavirus, HFRS is also affected by environmental and meteorological factors(15-22). Climatic and environmental anomalies directly contribute to the outbreak or expansion of various public health diseases, including HFRS(23). In Finland, Puumala hantavirus infection was not associated with cardiovascular disease, lung disease, kidney disease, and cancer but was associated with long-term smoking(24). Compared with our previous reports on HFRS(25), the present study (1) provides detailed patient information, (2) covers a long period from 2005 to 2019, (3) provides data visualization, and (4) analyzes the relative risk of diseases for different occupations, sexes, and ages.

## Materials and methods

### Data collection

Patient data were obtained from the Tai’an CDC. Every hospital in the Tai’an area reports cases to the Centers for Disease Prevention and Control (CDC) at all levels through the Infectious Disease Detection System. This system does not contain patient treatment information. All cases are clinically confirmed based on clinical manifestations and treatment reactions. However, it is not clear whether there is a virological diagnosis. Meteorological data from 2005 to 2018 were obtained from three meteorological monitoring stations around Tai’an. This study included the averaged data from these three monitoring sites. Population information was obtained from Tai’an Statistics Bureau. We received research approval from the Ethics Committee of Shandong First Medical University (ethics approval No: 202005004).). All patient data were anonymized.

### Methods and models

We calculated the numbers of cases in each year from 2005 to 2019 and plotted them on a density map. Similarly, we calculated the monthly numbers of cases and deaths over 15 years. We also plotted the age distributions of the patients on a density map. We calculated the relative risk (RR) for different occupations, age groups, and sexes. The population base of each subgroup was the average reported in the statistical yearbooks of Tai’an in 2011, 2012, and 2014. Considering the floating population every year, the total population of Tai’an is 5.6 million, of which the agricultural population accounts for approximately 50%. Since one patient in the 50–59 age group did not report his or her sex, we divided 1 equally between the male and female groups by 0.5. We used Joinpoint Trend Analysis version 4.5 to analyze the annual HFRS trends from 2005 to 2019. The principle of the software can be explained by referring to the following pages (https://surveillance.cancer.gov/joinpoint/). We used the “mgcv” software package (https://cran.r-project.org/web/packages/mgcv/index.html) to fit the generalized additive model (GAM) in which a linear predictor is given by the sum of the smoothing functions of user-specified covariates plus the normal parameter components of the generalized linear model.

We used the “dlnm” package (https://cran.rproject.org/web/packages/dlnm/index.html) to establish the distributed lag model of morbidity and meteorological factors. The first step involved selecting two sets of basic functions for exposure-lag-response correlation modeling. The cross-basis function generates the basic matrix of the exposure reaction and the lag-reaction relation and combines them to form the cross-base cross-basis by a special tensor product. We performed modeling using the “glm” function and forecasting with “crosspred” function. We divided each year into the first and second halves to perform seasonal analysis. We set the lag to 15, the temperature range to -12–33°C, and the relative humidity to 20–100%.

### Statistical analysis

The tests of significance used a Monte Carlo permutation method in Joinpoint Trend Analysis. Other analyses were mainly performed in R 3.61 (https://www.r-project.org/) language. All statistical tests with p <0.05 were considered statistically significant.

## Results

### Epidemic profile

From 2005 to 2019, 482 cases of HFRS occurred in the Tai’an area, corresponding to an average annual incidence rate in the population of 0.57/100,000. A total of nine patients died. The case fatality rate of HFRS was 1.87% (95% confidence interval CI: 0.66–3.08%). Eight (1.66%) cases were initially misdiagnosed with severe fever with thrombocytopenia syndrome (SFTS).

### Time distribution

Analysis of 15 years of HFRS cases between 2005 and 2019 showed two incidence peaks, in 2005–2006 and 2017–2019. These peaks accounted for 47.5% of all cases. The largest number of cases (n=60) occurred in 2005. The lowest number of cases (n=13) occurred in 2010 (Figure 1A).

**Figure 1.**
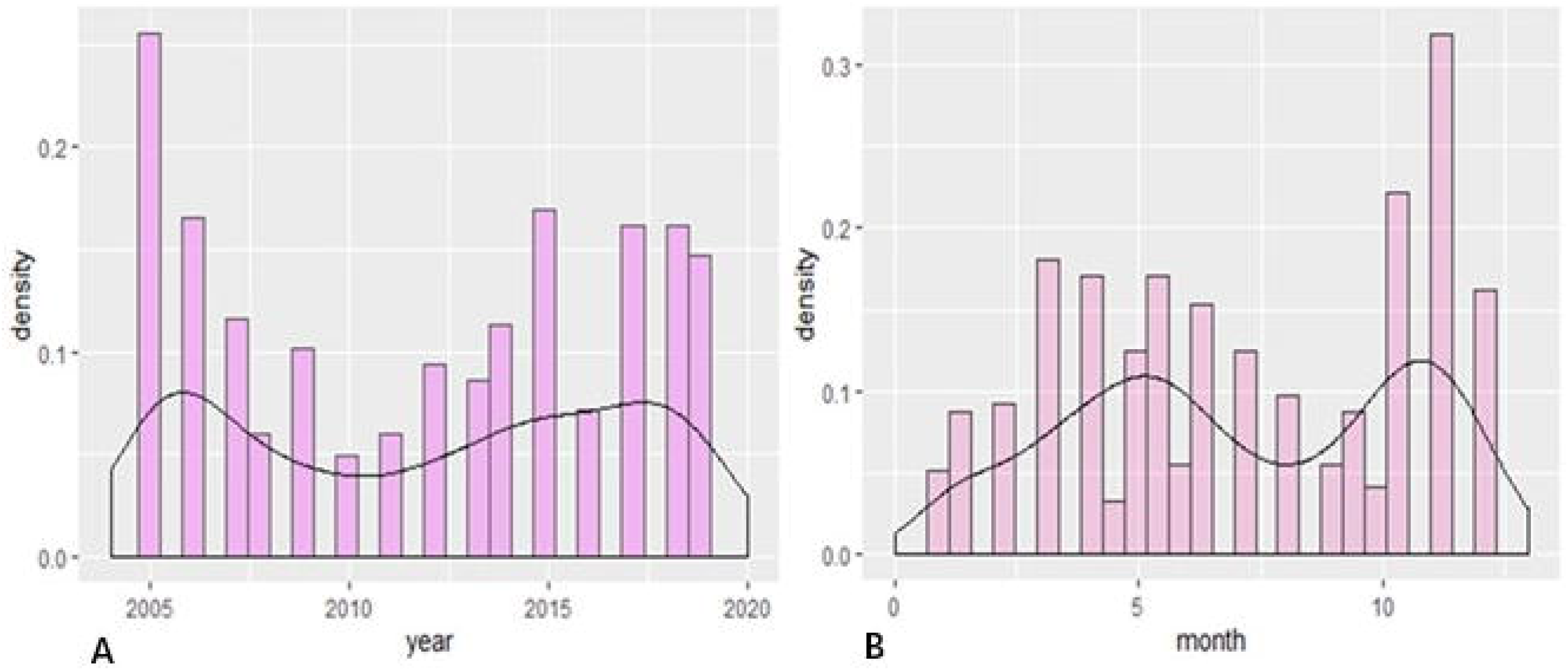
A. Distributions of the annual numbers of cases from 2005 to 2019. B. Distributions of the monthly numbers of cases from 2005 to 2019.

Exploration of the total numbers of cases per month over 15 years also showed two peaks of HFRS incidence in the Tai’an area, one in the summer months and the other in the winter months. The summer peak occurred from March 16 to June 15, with a total of 160 cases over 15 years (33.2%). The winter peak occurred from September 16 to December 15, with a total of 172 cases (35.7%) Figure 1B.

Among the nine deaths in 15 years, six occurred during winter months (November–January).

### Population distribution

#### Sex distribution

Over the 15-year study period the 142 female patients accounted for 29.5% of all cases while the 340 male patients accounted for 70.5% of patients. The average annual incidence rate in women was 0.35 per 100,000 population (95% CI: 0.13–0.57 per 100,000). The average annual incidence rate in men was 0.81 per 100,000 people (95% CI: 0.47–1.14 per 100,000).

#### Occupation distribution

The three occupations with the highest numbers of cases were farmers (74.07%); intellectuals, including teachers, students, cadres, medical staff, and retirees (12.03%); and workers (8.71%). Six patients were medical staff (1.24%).

#### Age distribution

The median patient age was 47 years (range: 3–85 years). The patients included 29 cases under 18 years (6.02%), 97 cases between 19 and 35 years (20.12%), and 296 cases between 36 and 65 years (61.41%). Only 60 patients were aged >65 years (12.45%). The peak age of onset for HFRS was between 36 and 65 years.

### Relative risks

#### Occupational risk

Compared with the whole population in the Tai’an area, the risks were significantly lower in young children, primary school students, and middle school students compared with the risk in the whole population (Figure 2). The risk of disease among college students, teachers, and non-rural medical staff was moderate with no significant difference. Farmers (25-64 years old) had a higher risk of the disease, with an RR of 2.00 (95% CI: 1.73–2.32, p <0.05). The rural medical staff had the highest risk of developing the disease, with an RR 5.05 (95% CI: 1.89–13.51, p <0.05).

**Figure 2.**
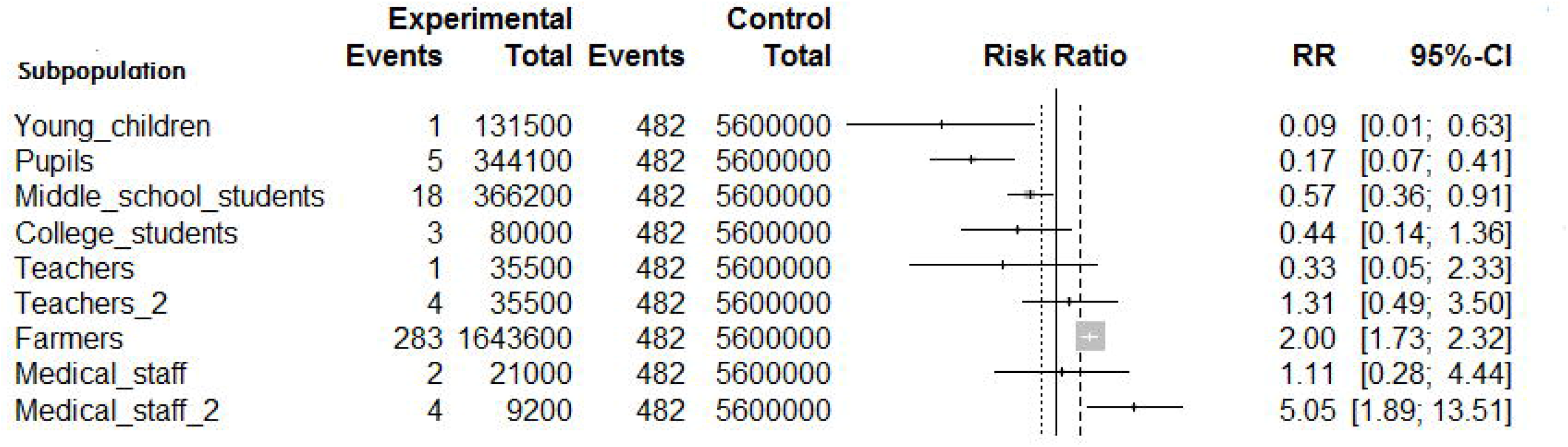
Risk of different populations compared with that of the whole population. Teachers: Teachers outside of rural areas. Teachers_2: Teachers in rural areas. Medical_ staff: Medical staff outside of rural areas. Medical_staff_2: Medical staff in rural areas.

#### Age and sex risks

Compared with the control group, women aged 0–39 years had a significantly low risk of onset (Figure 3). Women over 40 years of age had a moderate risk, with no significant difference. Men aged 0– 19 years had a low risk of the disease (p <0.05, Figure 3). However, men aged 30–79 years had a high risk of the disease. The risk was the highest for 50–59 years of age, with an RR of 2.16 (95% CI: 1.70–2.76, p <0.05). The risk of developing the disease in women aged 10–79 years was lower than that in men (p <0.05, S1 Fig).

**Figure 3.**
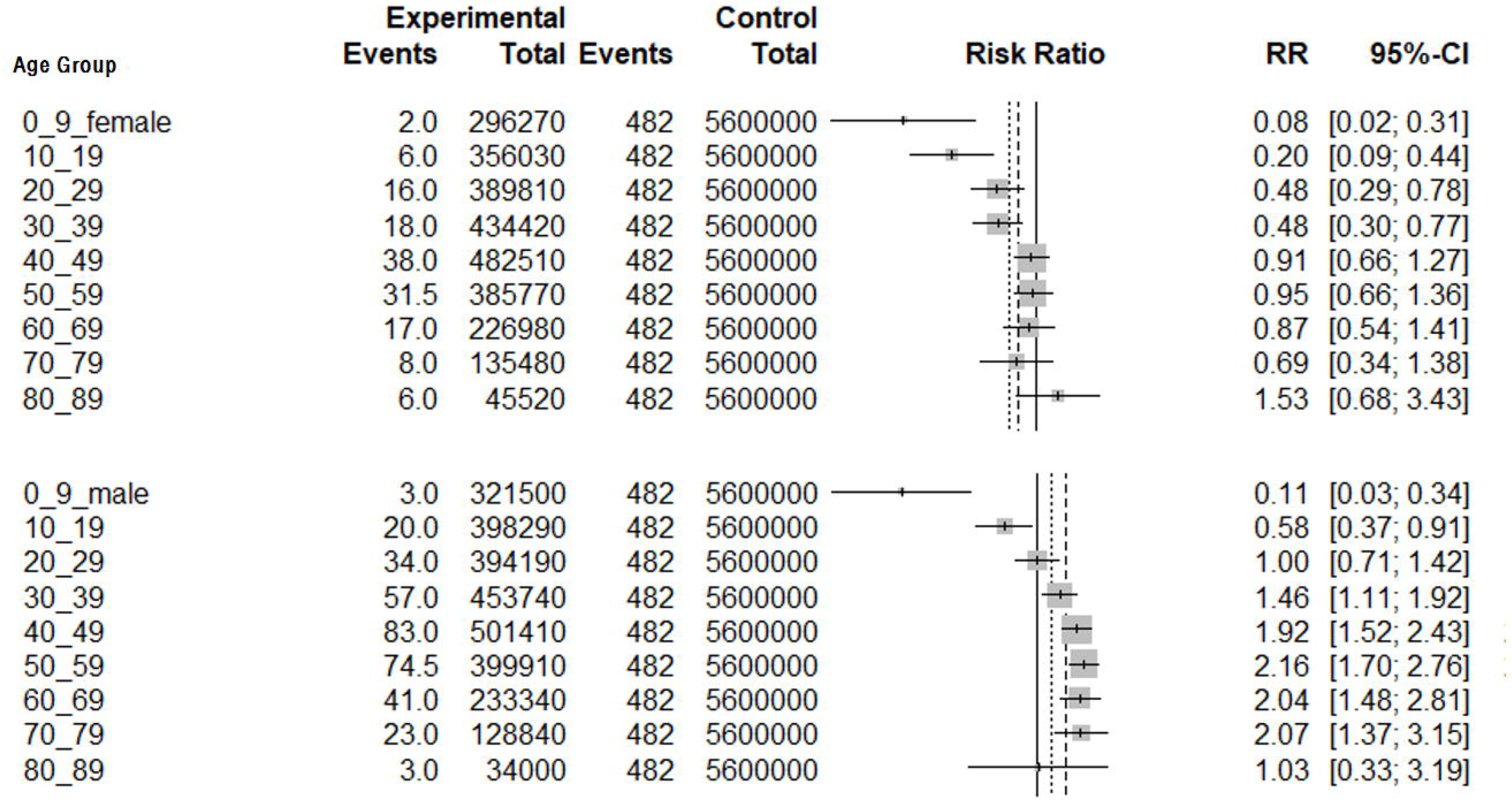
Risks of disease in different age groups compared to the whole population (women on top, men below).

#### Multiple joinpoint models

The joinpoint models showed that the total number of cases decreased by an average of 33.32% per year from 2005 to 2008 (p <0.05, Figure 4). From 2008 to 2019, the incidence increased by an average of 7.85% per year (p >0.05). For female patients, the incidence increased by an average of 0.93% per year from 2005 to 2019 (p >0.05, S2 Fig) and decreased by an average of 38.71% per year from 2005 to 2008 in male patients (p <0.05, S3 Fig). However, from 2008 to 2019, the incidence in male patients increased by an average of 9.24% per year (p <0.05).

**Figure 4.**
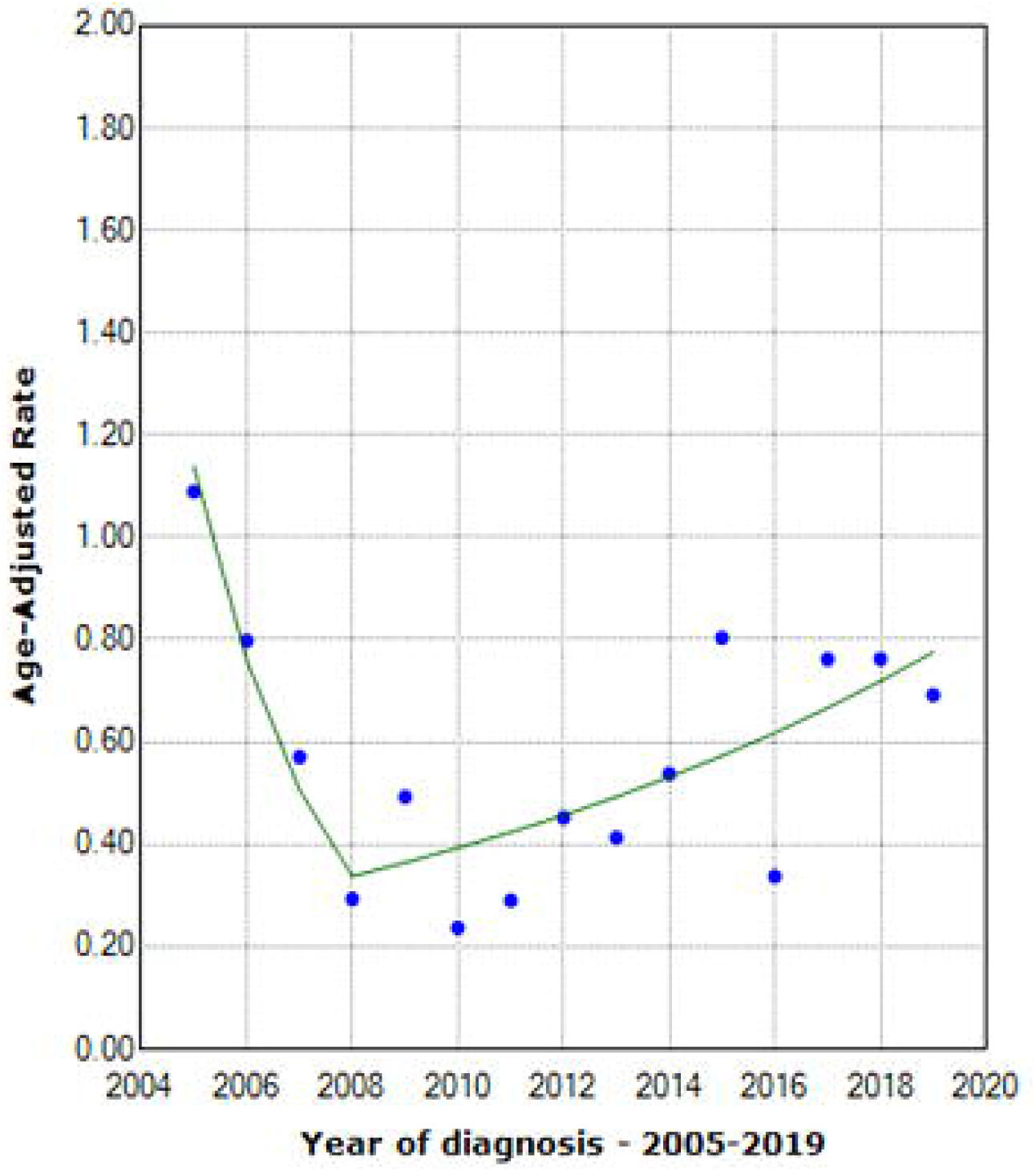
Joinpoint models showing the changes in average annual incidence rates from 2005 to 2019.

#### GAMs

We observed a positive correlation between temperature and disease risk from January to June (p <0.01, Figure 5A) but did not observe significant differences in risk for relative humidity (p=0.059) or sunshine duration (p=0.38). Overall, for every 1-unit increase in temperature, the risk of disease increased by 3%.

**Figure 5.**
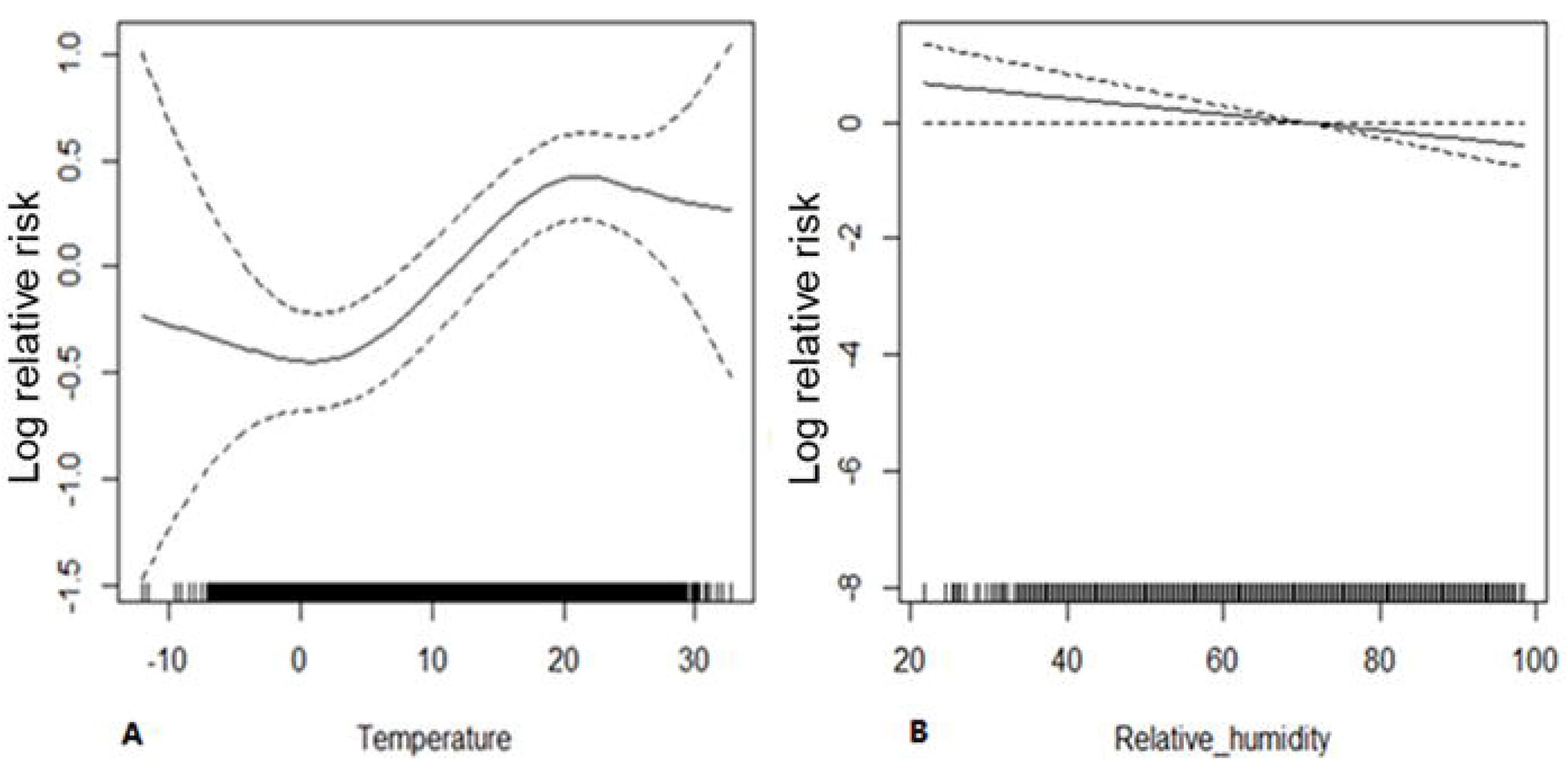
A. Relationship between temperature and disease risk. B. Relationship between relative humidity and disease risk.

However, we observed was a negative correlation between relative humidity and disease risk (p <0.05, Figure 5B) from July to December, while temperature (p=0.17) or sunshine hours (p=0.24) did not differ significantly. Overall, for every unit increase in relative humidity, the risk of disease decreased by 1%.

#### Distributed lag models

We built a bi-dimensional distributed lag nonlinear model (DLNM) using the DLNM package. We found that compared to the reference temperature (20°C), 1–2 lags had a low risk at low temperatures (<15°C) from January to June. However, at high temperatures (>30°C), compared to 20°C, 7–14 lags showed a lower risk (Figure 6A). From July to December, there was a high risk for 4–10 lags at low temperatures (<15°C) compared to 20°C. However, at low temperatures, 12 – 15 lags had a low risk. At high temperatures, 7–12 lags showed a low risk (Figure 6B).

**Figure 6.**
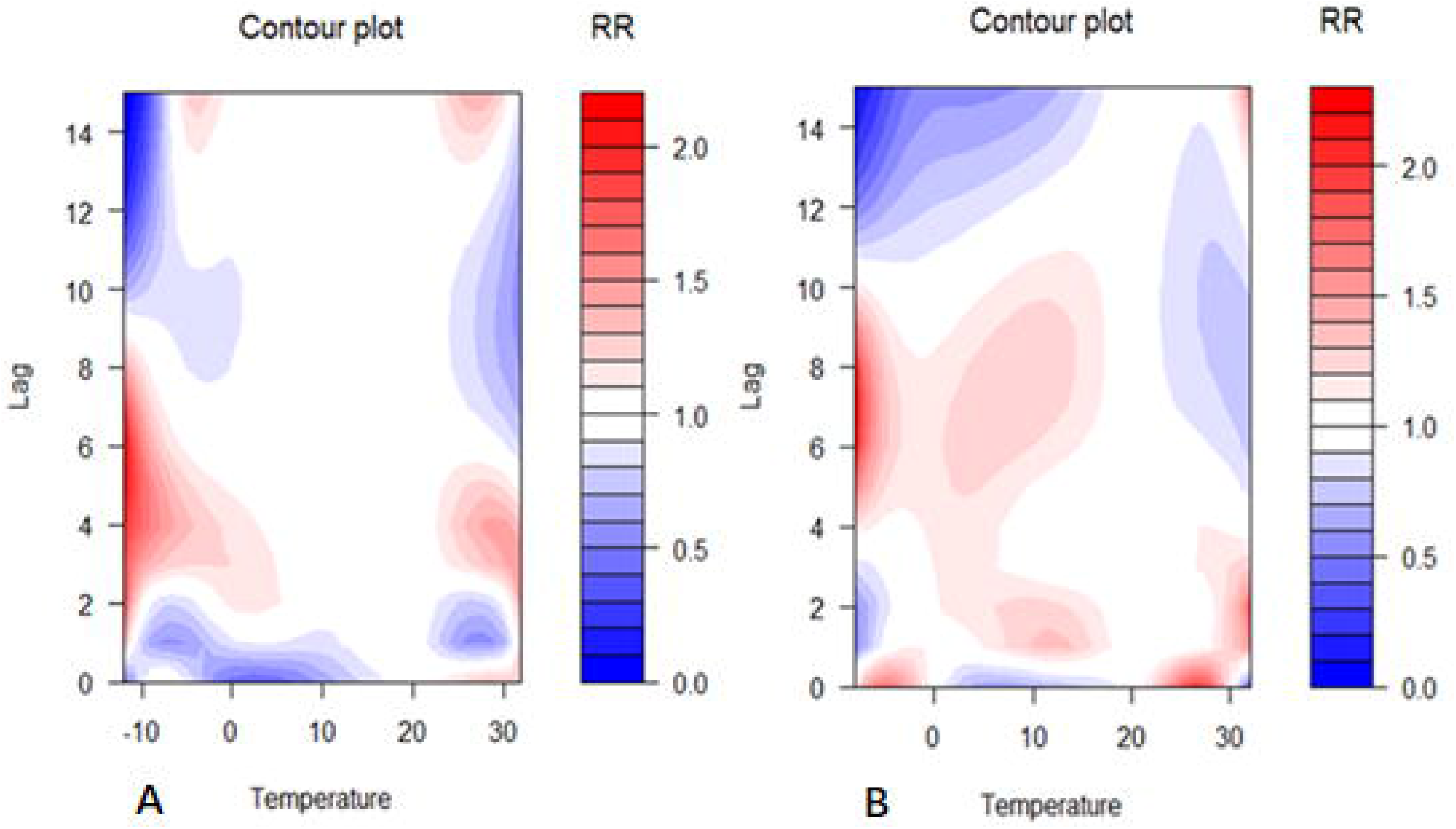
A. Heat map of the incidence risks and lags at different temperatures from January to June. B. Heat map of the incidence risks and lags at different temperatures from July to December.

From January to June, 1–14 lags showed a low risk of developing the disease at a low relative humidity (<40%). From July to December, when the relative humidity was low, there was a high risk compared to the control (relative humidity 60%) of 7–12 lags. When the relative humidity was very high, there was a low risk for 7–10 lags.

We also established distributed lag linear models (DLMs) for seasonal analysis to calculate cumulative disease risks. From January to June, at low temperatures, -5°C rose to 5°C, after 15 lags, the cumulative risk of disease increased (RR=2.4,95% CI: 1.48–3.88, p <0.05). At high temperatures, 15°C increased to 25°C and after 15 lags, the cumulative risk of RR=0.004 (95% CI: 0.001–0.014, p <0.05). In the same period, after 15 lags, when the relative humidity increased from 30% to 40%, the cumulative risk of RR was 0.93 (95% CI: 0.76–1.13, p >0.05). However, the cumulative risk of RR=0.63 (95% CI: 0.08–5.21, p >0.05) increased when the relative humidity increased from 80% to 90%.

From July to December, at low temperature, -5°C rose to 5°C, after 15 lags, the cumulative risk of RR=1.46 (95% CI: 0.86–2.46, p >0.05). At high temperatures, 15°C increased to 25°C, and after 15 lags, the cumulative risk of RR=0.004 (95% CI: 0.0001–0.28, p <0.05). In the same period, after 15 lags, when the relative humidity increased from 30% to 40%, the cumulative risk RR was 0.66 (95% CI: 0.53–0.83, p <0.05, Figure 7A&B). Under the same conditions, the cumulative risk of RR=0.14 (95% CI: 0.05–0.38, p <0.05) increased when the relative humidity increased from 80% to 90%.

**Figure 7.**
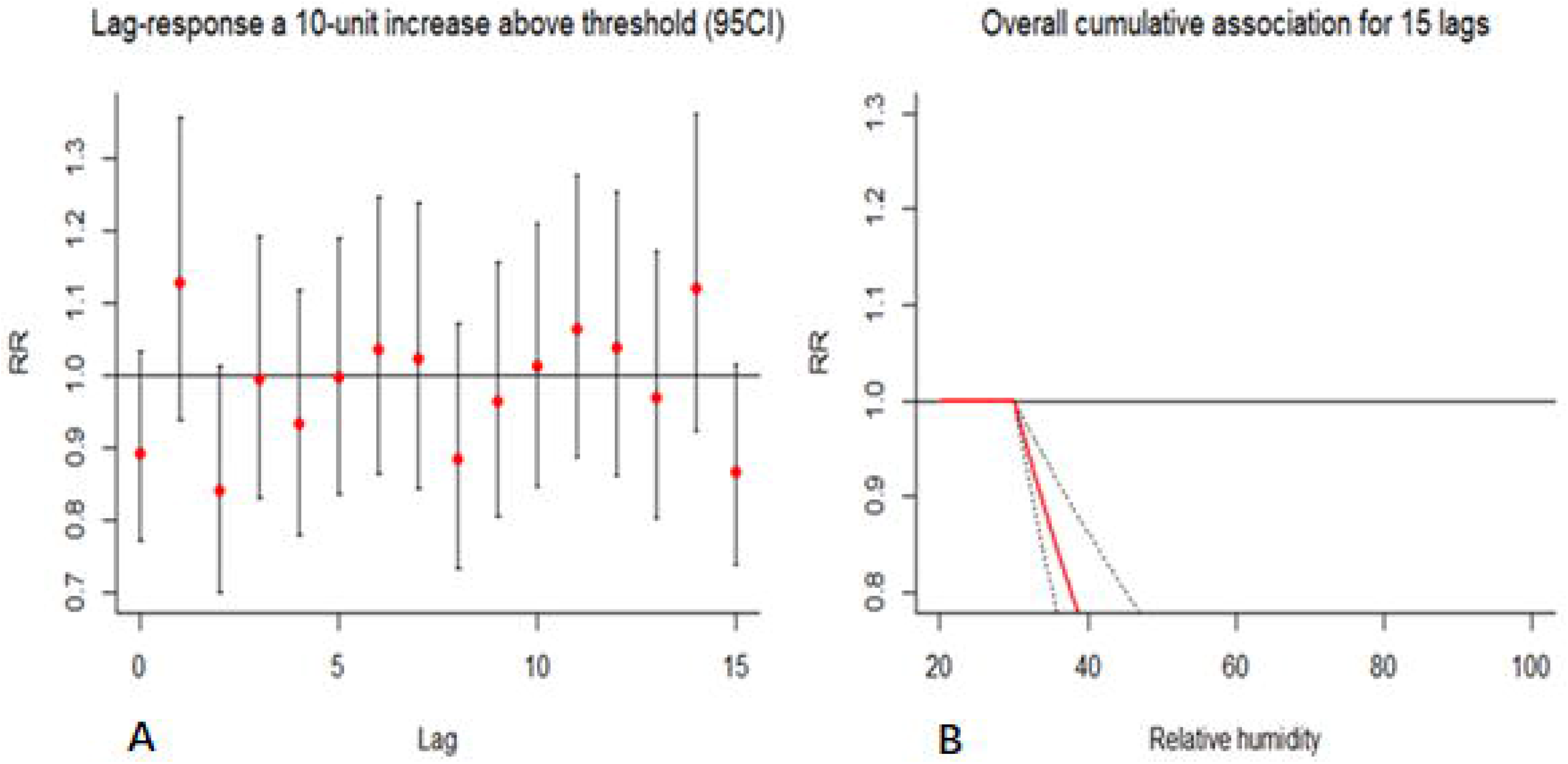
A. Relative humidity increases by 10 units (30%–40%) and the risks associated with different lags. B. Relative humidity increases by 10 units (30%–40%) after a lag of 15 units on the cumulative risk of disease.

## Discussion

Together with the CDC in Tai’an City, this study conducted a comprehensive analysis of HFRS over 15 years. The average annual incidence of the disease was 0.57/100,000, and 1.66% of cases were initially misdiagnosed as SFTS. Two incidence peaks were observed in the past 15 years. In terms of months, there was a summer peak and a winter peak. In this study, 70.5% of the patients were male. Regarding occupation, 74.07% of cases were farmers and migrant workers. Four patients were rural medical staff. From the perspective of profession, compared to the whole population, the RR of incidence for the agricultural population was 2.00. The incidence risk RR of rural medical staff was as high as 5.05. In terms of age, men aged 30–79 years were at higher risk. In terms of sex, the risk of disease in women aged 10–79 years was lower than that in men. Overall, the incidence rate decreased from 2005–2008. In the first half of the year, for every unit increase in temperature, the risk of disease increased by 3%. In the second half of the year, for every unit increase in relative humidity, the risk of disease decreased by 1%. From January to June, the temperature increased from -5°C to 5°C and the cumulative risk of disease increased after 15 lags (RR=2.4). However, we found that the cumulative risk was reduced at high temperatures. Specifically, when the temperature rose from 15°C to 25°C, the cumulative risk decreased after 15 lags (RR=0.004). From July to December, after 15 lags, when the relative humidity increased from 30% to 40%, the cumulative risk RR was 0.66. The cumulative risk (RR=0.14) increased when the relative humidity increased from 80% to 90%.

Our research showed that the predominant age groups in our local patients were middle-aged. Furthermore, most patients were men and the main occupation was farming, consistent with the findings of previous reports(1, 13, 14, 26). The risk prediction model showed that cultivated land had the highest risk factor for HFRS(15). In contrast to the previous literature(27), the disease showed a bimodal structure in this region throughout the year, rather than a single peak. These double peaks may be related to the living habits of local mice.

To our knowledge, this study is the first to report the relative risks of different groups. Previous studies have suggested that farmers account for the majority of patients but do not indicate farmers have the highest risk of disease. Our study found that the risk of disease in farmers was twice as that in the whole region. On the one hand, farmers’ income is low, and their living environment is poor, and they may thus have more opportunities to come into close contact with rats; on the other hand, in agricultural production, farmers have more direct or indirect contact with farm voles, which increases their risk of infection. It is concerning that the RR of infection of medical staff in rural areas was 5.05. However, the risk of infection was not high for medical staff outside of rural areas. The income of rural teachers is similar to that of rural doctors, but the risk for rural teachers is not as high as that of rural doctors. Medical workers in rural areas are in contact with many patients with fever of unknown causes or with hidden infections, which are difficult to detect. This may be the reason the risk in rural medical staff was higher. In short, from the high risk in rural medical workers, we highly suspect the possibility of human-to-human transmission, although these people may also be infected at home. Because the number of infections was limited, it is worthy of paying attention to the rural medical staff and using statistical analyses of their past diseases over a larger area to put forth a stringent argument. The risk of infection in men aged 30–79 years was higher than that of the whole population and for women in the same age group. Men in this age group have a higher risk of disease, which may be related to their patterns of physical activity and possibly to differential susceptibility according to sex.

Previous studies have used different mathematical models to predict HFRS, including the ARIMA model(28). Our study used four models to fit the data and predict the analysis. The multiple joinpoint models showed that the total number of cases decreased by an average of 33.32% per year from 2005 to 2008. This finding may be because of economic development, rural urbanization, or improved living conditions. We have observed a slight increase in the number of cases in recent years. In China, significant positive and negative correlations between HFRS incidence and urbanization have been reported in the primary (1963–1990) and secondary (1991–2010) stages of urbanization, respectively(29).

Although previous studies reported a relationship between HFRS and rainfall(30, 31), our analysis at different monitoring points showed wide rainfall variations between the meteorological monitoring points about 100 km apart; thus, we felt that the error in including rainfall in the model was too large. Therefore, we do not advocate the use of rainfall to predict the incidence of the disease. The GAM showed that the risk increased by 3% for every 1 unit increase in temperature in the first half of the year and decreased by 1% for every 1 unit increase in relative humidity in the second half of the year. Meteorological factors have a lag effect on disease onset (27). From January to June, at low temperature (−5–5°C), after 15 lags, the cumulative risk was significantly higher. However, the cumulative risk was reduced at high temperatures. From July to December, the cumulative risk of 15 lags decreased with an increase in relative humidity at different stages. This shows that air temperature and relative humidity are important environmental factors affecting HFRS.

## Conclusions

The incidence of HFRS in the Tai’an area of Shandong Province showed double peaks every year. The summer peak occurred from March 16 to June 15, while the winter peak occurred from September 16 to December 15. Compared with the whole population, the RR value of disease risk of farmers is 2.00. Rural medical staff had the highest risk of infection, with an RR as high as 5.05. It is worthy of exploring the possibility of human-to-human transmission of HFRS among rural medical workers in the future. The risk of infection in men aged 30–79 years was higher than that of women in the same age group and the population as a whole, which may be related to a wide range of activities, more participation in agricultural production activities, and sex susceptibility of men. With economic development, the incidence of the disease remains low. From January to June, the temperature increased from -5°C to 5°C and, after 15 lags, the cumulative RR was 2.4. The increase from 15°C to 25°C and after 15 lags, showed a cumulative RR of 0.004. From July to December, after 15 lags, the relative humidity increased from 30% to 40%, with a cumulative risk RR of 0.66. The cumulative RR was 0.14 when the relative humidity increased from 80% to 90%. Temperature and relative humidity were the two main environmental factors affecting the incidence of hemorrhagic fever.

## Data Availability

All the data can be found in the article.

## Notes on contributors

Z.Y., conceived and designed the study. X.B., S.Y, A.Z., and Z.Z., processed the data. Y.L., T.W., and C.Z., performed statistical analysis. X.B., S.Y., A.Z., Z.Z., and Z.Y., contributed to establishing the mathematical models. X.B., and Z.Y., completed the writing of the manuscript.

## Acknowledgements

We thank the Tai’an CDC for providing case data and an English editing company for language proofreading.

## Supporting information

S1.Fig Risks for different age groups according to sex.

S2.Fig Joinpoint models showing the changes in average annual incidence rates of females from 2005 to 2019.

S3.Fig Joinpoint models showing the changes in average annual incidence rates of males from 2005 to 2019.

